# Predictors of Anemia in Ethiopia: A Systematic-Review of Machine Learning Approaches

**DOI:** 10.1101/2025.05.08.25327246

**Authors:** Mahider Ayalew, Tadesse Mamo, Makda Abate Belew, Agmasie Damtew Wale

## Abstract

**Background:** Anemia remains a critical public health issue globally which is disproportionately affecting population in low- and middle income countries, with sub-Saharan Africa particularly Ethiopia experiencing high prevalence rates, Despite ongoing interventions, understanding the multifactorial causes of anemia and enhancing predictive Modelling through modern analytic approaches remains limited.

**Objective:** This systematic review aims to evaluate and synthesize current evidence on the application of machine learning algorithm for predicting anemia among various population in Ethiopia, focusing on identifying predictive models used, key predictors and methodological strength of those existing studies.

**Methods:** Following the PRISMA 2020 guidelines, a comprehensive search was conducted across PubMed, science Direct, HINARI, and Google Scholar from October 25 to November 10 2024. Observational studies employing Machine learning algorithm to predict anemia in Ethiopia were included. The quality of included studies was evaluated using BSA Medical Sociology Group Assessment Tool.

**Results:** Out of 513 initially retrieved records, four studies met the inclusion criteria. These Studies targeted Children under five, pregnant women, and young girls, Utilizing algorithms such as Random Forest, Logistic Regression and Boruta algorithm .the random Forest model emerged as the most frequently and effectively used technique due to its Robustness and capacity for handling complex data. Anemia prevalence across the included studies ranged from 26% to 57%. A total of 28 unique predictor variables were identified.

**Conclusion and Recommendation:** Machine learning algorithms, particularly random forest, offer promising tools for accurately predicting anemia in Ethiopia by integrating wide range of socio-demographic and health-related factors, however, the limited number of studies and population specific focus highlight the need for more comprehensive and generalizable research to inform effective public health intervention.

## Introduction

Anemia is a significant global health issue, affecting nearly one-quarter of the world’s population, particularly in low- and middle-income countries. According to the World Health Organization (WHO), anemia poses a serious threat to public health and is a major contributor to maternal and child morbidity and mortality(1).The burden of anemia is especially pronounced in Africa, where factors such as poverty, inadequate nutrition, infectious diseases, and limited access to healthcare exacerbate the prevalence of this condition(2).

In Africa, the prevalence of anemia varies widely, with certain regions experiencing rates as high as 50%(3). This discrepancy highlights the need for targeted interventions and effective monitoring systems. Within the continent, Ethiopia stands out as a country facing a significant challenge regarding anemia, with estimates indicating that nearly half of all children under five and a substantial percentage of women of reproductive age are affected(4). Specifically, the national prevalence of anemia in Ethiopia is estimated to be 56% among children under five years old and 23% among women of reproductive age(3).This presents a critical public health challenge that requires urgent attention, as anemia is linked to increased risks of maternal and child morbidity and mortality(4).

Despite the efforts made to combat anemia in Ethiopia, gaps remain in understanding its multifaceted causes and the effectiveness of current interventions. Traditional methods of anemia prediction often rely on demographic and clinical data, which may not capture the complex interactions between various risk factors (5). Machine learning approaches offer a promising alternative by leveraging large datasets to identify patterns and predict outcomes with greater accuracy. However, there is limited systematic research on the application of these methods specifically for predicting anemia in the Ethiopian context (3, 6).

The challenge lies in the lack of comprehensive studies that synthesize existing machine learning approaches tailored to anemia prediction in Ethiopia. While numerous studies have explored anemia in various populations, few have systematically reviewed the efficacy of machine learning techniques in this area. For instance, while studies have employed machine learning algorithms to predict anemia among youth girls and under-five children in Ethiopia, they often focus on specific populations without a broader synthesis of findings or methodologies (5–7). This gap hinders the development of effective predictive models that could inform public health strategies and interventions tailored to the Ethiopian population.

In light of these challenges, this comprehensive systematic review aims to evaluate and synthesize the current state of machine learning applications for predicting anemia, with a focus on their utility within Ethiopia. By addressing this gap, the study seeks to provide valuable insights that can enhance predictive accuracy, guide policy decisions, and ultimately contribute to reducing the burden of anemia in Ethiopia.

## Methods and materials

### Eligibility criteria

To declare the inclusion and exclusion criteria in this systematic review and meta-analysis the researchers followed PICO and Study design Filter Approach mainly CoCoPop (condition, context, and population Questions)

### Inclusion criteria’s

#### Setting

Ethiopia, both rural and urban, community

#### Study design and period

This systematic review and meta-analysis encompass all observational study designs that focus on predicting anemia in Ethiopia using a machine learning Approach.

#### Participants

All populations (Women, youth, Children, adolescents, Adults)

#### Exclusion criteria

Review and qualitative studies, articles with unknown primary outcomes.

#### Search strategy

The objective of this review was to systematically Review articles done on the prediction of Anemia among Different Populations in Ethiopia using Machine Learning Approach. We followed the PRISMA-2020 protocol for this systematic review (7). Three authors, namely, MA, AD, and MA conducted a comprehensive search across multiple databases, including PubMed/Medline, HINARI, Science Direct, and Google Scholar. The review was conducted between October 25th and November 10th, 2024. To ensure there was no duplication, all retrieved materials were imported into Endnote X20. The search term used in the databases was “ Employing AND Machine Learning AND predictors AND Anemia AND Ethiopia” as the MESH term. The protocol for this review is under process of registration on the International Prospective Register of Systematic Reviews (PROSPERO).

#### Study selection

Following a systematic search across all relevant databases, duplicate studies were excluded. Studies that were deemed unrelated based on their titles were also excluded. The remaining studies underwent a thorough review of their abstracts and full texts by two independent reviewers, MA and AD. In cases where there was disagreement between the reviewers, discussions were conducted to reach a consensus. If a consensus could not be reached, a third reviewer, MA, made the final decision on which articles to include in the review. Only studies with appropriate study methodologies were included for full-text review **(Fig1**).

**Figure 1.**
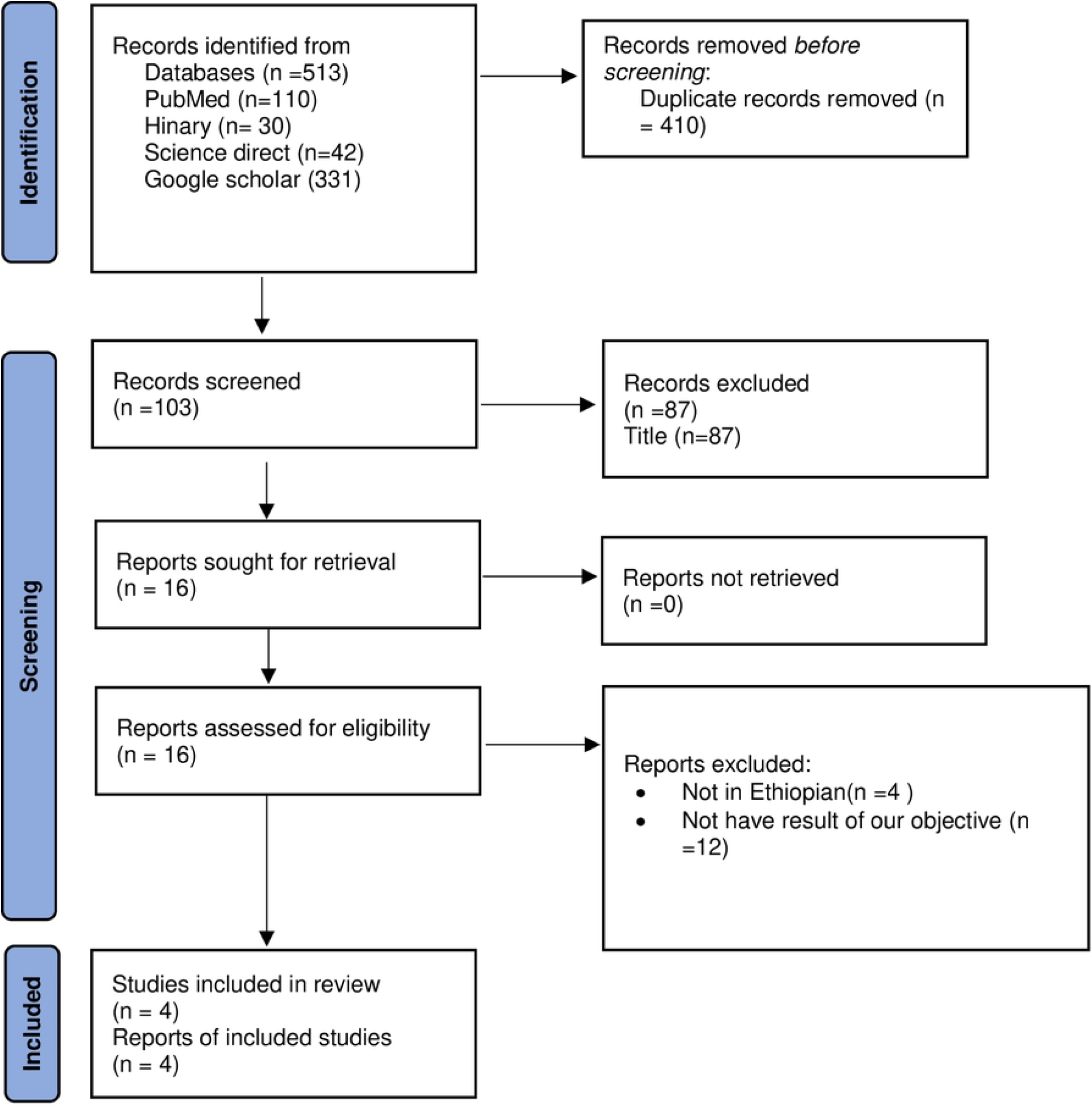
PRISMA Flow chart for the study selection process

#### Data extraction

Two independent investigators, M.A and MA performed the data extraction from the selected studies. A custom “Data Extraction Form” was thoughtfully created using Microsoft Excel 2013 to facilitate a robust and standardized process for extracting relevant data from the eligible studies included in this meta-analysis. The data extraction form included relevant information such as outcome measures, including reported estimates or sufficient data to calculate the pooled prevalence of Intimate partner violence against women in Ethiopia, as well as odds ratios (ORs) with 95% confidence intervals (CIs). Publication details, such as the title, author, publication status (published or unpublished), journal name, year of publication, study period, and the region where the study was conducted, were also recorded. Additionally, the form captured study design details and participant information, including the number of individuals involved and population characteristics such as age, educational status of women, educational status of partner, occupation, polygamy practice, autonomy on decision making, history of abortion.

#### Assessment of the methodological quality of included studies

Two reviewers independently assessed each study’s scientific strength and quality using the BSA Medical Sociology Group Quality Assessment Tool. The criteria have seven quality indicators that assess the three levels of bias: 1-2 (low), 3-5 (moderate), and 6-7 (high). (1) an appropriate research design; (2) an appropriate recruitment strategy; (3) a reported response rate; (4) a sample representative of a similar population; (5) objective and reliable measures used; and (6) an appropriate sampling strategy are the seven quality indicators to be evaluated. Power Calculation and Justification of Numbers Reported (7) Appropriate Statistical Analysis.

#### Data extraction and analysis

The data from the included studies were compiled into a Microsoft Excel spreadsheet by three authors, and the accuracy was checked by the remaining researchers. For each study, the initial author’s name, the year it was published, types of research, sample size, sampling technique, study population, data collection techniques, and the study’s design were all gathered. Parameters were also retrieved, and discrepancies amongst data extractors have been resolved through discussion with the authors, via the PRISMA standard. Moreover, the article is moved to the “Resolve disputes” list if the authors cannot agree on whether or not to include the study or the justification provided for its exclusion.

In this study, a meta-analysis of the finding was not conducted due to differences in methods and participant types between studies. For instance, this study used both quantitative and qualitative methods to investigate the elements that make acceptance of e-health technology. The study’s participants also varied (women, children, and Adolescents). Moreover, some studies’ statistical interpretations that supported the conclusions were not practicable. Accordingly, the current study adopted the analytical approach used in earlier systematic reviews. In this way, the facilitators were examined based on how frequently they appeared in the studies and the investigation of this approach resulted in accurate findings.

## RESULT

### Search results

A total of 513 articles were obtained up on initial searching from PubMed, Science Direct, HINARI and Google Scholar. A total of 410 articles were removed due to duplications. Finally a total of 87 articles were excluded by observing their title. Consequently, only 16 articles were subject to a full-text review. Finally, 4 articles were selected to be included in our review **(fig1)**.

### Characteristics of studies included in this review

In our review the filtered articles were studies done on predictors of anemia that use only machine learning algorithm approach. Articles included in this study were conducted among, pregnant women, children age of <5 and young Girls health care workers and community. The majority of the participant were male in the two articles(6, 8). Whereas as remaining two articles were only done among female populations (9, 10) and regarding residence of participant’s majority were from rural area (6, 8–10). Furthermore from all two of the studies predict using random forest Model (9, 10)(Table 1)

**Table 1.** Summary of baseline characteristics of the articles that were previously published and included studies in the systematic review, 2024.

| Author<br>&Year of<br>publication | Population | Total<br>sample | Predictive<br>Model used | Prediction<br>model<br>accuracy | Quality<br>score | Gender<br>(%) | Resident |  |  |
| --- | --- | --- | --- | --- | --- | --- | --- | --- | --- |
|  |  |  |  |  |  | female | Male | Urban | Rural |
| Tesfaye et al<br>(2024) | children<5 | 8,482 | Logistic<br>regression | 66% | High | 4084 | 4398 | 861 | 7621 |
| Dejene et al<br>(2022) | pregnant<br>women | 11,174 | Random<br>Forest | 91.34% | High |  |  |  |  |
| Abdulaziz.<br>K(2023) | children<5 | 9,501 | Boruta<br>algorithm | NA | High | 4855 | 1805 | 1805 | 7696 |
| Alemu<br>.B(2024) | young<br>girls | 5,642 | random<br>forest | 82% | High |  |  | 2002 | 3640 |

### Prevalence of the Anemia

Prevalence of Anemia were reported in three (Tesfaye et al, Abdulaziz. K, Alemu. B) of articles included in our review according to the study of Tesfaye et al the prevalence of anemia among children under age of five were 57 %((6)and study of Abdulaziz. K’s study which were done among under five children it was 40%(8) and study done by Alemu. B among young girls the prevalence were 26%(10)(Table2).

**Table 2.** Summary of included studies on Prevalence and predictors of Anemia studies done using machine learning approach, in Ethiopia, 2024.

| Author | Anemia | Predictors |
| --- | --- | --- |
| Tesfaye et al | 57% | • Child age in months<br>• Age of mother<br>• Place of residence<br>• Wealth quintile<br>• Mother's education<br>• Employment status of mother<br>• Stunting<br>• Wasting<br>• Child morbidity<br>• Maternal anemia |
| Dejene et al |  | • duration of current pregnancy<br>• age<br>• source of drinking water<br>• history of contraceptive use<br>• respondent's occupation<br>• number of household members |
| Abdulaziz. K | 40% | • number of children<br>• distance to health facilities<br>• health insurance coverage<br>• residence<br>• mothers' wealth index<br>• type of cooking fuel<br>• number of family members<br>• mothers' educational status<br>• receiving rotavirus vaccine |
| Alemu .B | 26% | • Region<br>• wealth index<br>• Educational status<br>• Family size<br>• Marital status<br>• Source of drinking water<br>• Types of toilet facility<br>• Altitude<br>• Occupational status<br>• Media exposure<br>• Nutritional status |
|  |  | • Contraceptive method |

### Factors that Are Predictors of anemia

In this systematic review, a Total of 28 Predictor variables have been found in the four studies. The identified predictors of anemia from studies done using machine learning approach were Child age in months, Age of mother, Place of residence, Wealth quintile, Mother’s education, Employment status of mother, Stunting, Wasting, Child morbidity, Maternal anemia, number of household members, number of children, distance to health facilities, health insurance coverage, type of cooking fuel, receiving rotavirus vaccine, Region, Marital status, Source of drinking water, Types of toilet facility, Altitude, Occupational status, Media exposure, Nutritional status, Contraceptive method. (table2).

The predictors of anemia identified in the systematic review are categorized into socio-economic, demographic, and health-related factors, with specific studies highlighting each element.

Socio-economic factors include wealth quintile and mother’s education, both identified in Tesfaye et al(6), which indicate how economic status and educational attainment influence health outcomes. Employment status of the mother, highlighted in Tesfaye et al(6) and Abdulaziz K(8)., reflects the impact of job stability on access to resources. Additionally, the occupation of the respondent noted in Dejene et al(9) and household size discussed in Abdulaziz K(8). Illustrate how family dynamics affect resource availability. The type of cooking fuel, emphasized in Abdulaziz K(8).along with health insurance coverage, also from the same study, further underscore the socio-economic determinants of anemia.

Demographic factors include child age in months and maternal age, both highlighted by Tesfaye et al(6), illustrating the vulnerability of younger children and the influence of maternal age on child health. The place of residence, as identified in Tesfaye et al(6) and Abdulaziz K(8)., indicates disparities in health access between urban and rural areas. Region, mentioned in Alemu B(9)., reflects geographic variations in health outcomes. Marital status, discussed in Alemu B., and the number of children, noted in Abdulaziz K(8)., show how family structure can impact health resource allocation.

Health-related factors encompass maternal anemia, highlighted in Tesfaye et al(6), which has direct implications for child health, and child morbidity, also noted in Tesfaye et al(6), which can lead to nutritional deficits. Stunting and wasting, identified in Tesfaye et al(6), serve as indicators of malnutrition predisposing children to anemia. Nutritional status is a critical factor across studies, while the source of drinking water, noted in Dejene et al and Abdulaziz K(8)., underscores the importance of clean water for preventing disease. Types of toilet facilities, emphasized in Alemu B(9)., relate to sanitation and health outcomes. Finally, altitude, discussed in Alemu B(9)., and receiving the rotavirus vaccine, noted in Abdulaziz K(8). Highlight additional health determinants that can influence anemia prevalence.

## Discussion

The findings from our review highlight critical trends in studies focusing on predictors of anemia using machine learning algorithms, particularly among vulnerable populations such as pregnant women, children under five, and young females in Ethiopia.

The findings of this systematic review indicate that the random Forest Model was the Most Frequently utilized prediction model for selecting predictors of Anemia across the reviewed studies .this preference can be attribute to several key advantages associate with random forest algorithm, making it a robust and reliable choice for feature selection and predictive modeling.

One of the primary reason for the adoption of random forest is its ability to handle high – dimensional data effectively .unlike traditional statistical methods, random forest can manage large data set with numerous predictors without overfitting .it can enhance model stability and generalization(11) .additionally, random forest provides an inherent mechanism for feature importance ranking researchers to identify the most relevant predictors with high accuracy, which is beneficial in studies where selecting the optimal set of predictors is essential for improving model performance and interpretability .compared to other feature selection methods this random forest mitigates the risk of collinearity and bias in variable selection which makes it preferred choice algorithm (12, 13)

Furthermore, its non-parametric nature random forest to accommodate various types of data, including continuous, categorical, and missing values. Despite these advantage one drawback of random forest algorithm is its computational complexity, particularly when dealing with very large data sets, additionally while it provides insights into feature importance(14, 15)

### Socio-economic factors

Anemia-related health outcomes are revealed to be significantly influenced by mothers’ educational attainment and wealth quantile. Numerous studies have demonstrated a direct correlation between anemia and socioeconomic level. According to an Indian study, nations with higher socioeconomic position have lower rates of anemia than those with lower socioeconomic status. Two socioeconomic factors that influence children’s anemia are women’s wealth and health insurance coverage, according to a prior Ethiopian study (8). Children from the wealthiest home had a 48.3% prevalence of anemia, whereas children from the lowest household had a 68.1% prevalence, according to another Ethiopian study(6). According to the systematic review and meta-analysis study, children in Ethiopian low-socioeconomic families had a greater anemia status (16, 17)

Other socioeconomic characteristics that have been identified to be significant in various research reviews include occupation, mother’s lack of education, household size, kind of cooking fuel, and health insurance coverage (5, 18–22)

### Demographic factors

This review identified demographic factors like age of the child, maternal age, place of residence where they live In rural or urban settings, marital status and number of children as a demographic determinants of child hood anemia so, for instance, child age of 6 – 23 months of age are more likely to have child hood anemia than 24-59 months of age groups in previously conducted research using machine learning approach in Ethiopian context (6, 23–28)

On the other hand, maternal age is found to be another determinant for the prediction of childhood anemia in this review and other similar studies conducted In Ethiopia the maternal age particularly when the maternal age less than 20 it is found to be associated with childhood anemia (6) In one study which conducted in 55 lower and middle income countries it was found that early maternal age were related to the unexpected birth outcome including childhood anemia hence, increasing maternal age during pregnancy might have an added advantage towards good birth outcome in least developed countries where there are many teen age and adolescent pregnancy childhood anemia is associated with the maternal age when compared to late 20s those mothers in early 20s are more likely to have poor child birth outcomes like anemia (29) another study in Nigeria reported that when the mothers age increases the occurrence of anemia in their child decreases it revealed that Children whose moms were between the ages of 15 and 34 had the highest prevalence of anemia (73.5%).surprisingly, the prevalence of anemia declined with the mother’s age group. Mothers who gave birth to their first child between the ages of 10 and 19 accounted for 53.1% of all children between the ages of 6 and 59 months. Of these children, 72.5% were anemic; in contrast, only 54.7% of children born to moms who were at least 30 years old were anemic maternal age were associated with childhood anemia in another Bangladesh and Indian study (30) (23, 26, 28, 31–34).

Place of living is another predictor variable for children anemia. In our review, it was found to be one of the model’s consistent predictors. Other studies have found a substantial correlation between the place of residence and the outcome variable, childhood anemia. Accordingly, an Indian study found that children in rural areas are more prone than those in urban areas to suffer from severe and moderate childhood anemia (23) other nationally representative survey and systematic review studies also reported that place of residence is highly linked to the childhood anemia (26–28, 31, 35–38) the reason might be due to the disparity in health access in rural and urban areas.

Another predictor of childhood anemia using a machine learning prediction approach is marital status. In a similar vein, further research shown that this characteristic consistently predicts childhood anemia in various locations (32, 39, 40).Family structure and marital status are found to be the determinant factors of the childhood anemia in Indian and Mexico study (41, 42)

### Health-related factors

Children who are stunted and wasting are particularly vulnerable to childhood anemia due to their low immunity and the nature of malnutrition, which is one of the primary causes of childhood disease and mortality. Stunting and wasting are the two nutritionally related predictors of childhood anemia (24, 43–45).

The source of drinking water is another determinant for childhood anemia those children’s who have no access to safe and clean drinking water are more likely to had anemia when compared to the children’s who can access safe and clean drinking water (8, 46–49)

The types of toilet facilities are the other types of determinants for childhood anemia and including environmental factors like altitude (50, 51)

### Conclusion

Anemia remains a significant public health challenge in Ethiopia, Particular among children under five, pregnant women, and young girls. The systematic review high lights the effectiveness of machine learning approaches, particularly the random forest model, in predicting anemia and identifying those predictors, including maternal education, wealth status, child age, maternal age, and nutritional status, play a critical role in determining anemia prevalence in Ethiopia.

Despite advancements in prediction modelling, gaps remain in synthesizing findings across different populations and improving the generalizability of machine learning models for anemia prediction in Ethiopia. And as implication future research should focus on integrating more comprehensive datasets, improving model interpretability, and leveraging machine learning to develop targeted interventions. Addressing anemia though data driven approaches can enhance public health strategies and contribute to reducing its burden in Ethiopia.

## Data Availability

All relevant data are within the manuscript and its Supporting Information files.

## Declarations Acknowledgement

None

## Availability of data and materials

The corresponding author can provide the datasets used and/or analyzed in the current study upon reasonable request

## Authors contribution

All authors are participated in searching, screening, data extraction and writing up of the manuscript. All authors reviewed and approved the manuscript

## Ethics approval and consent to participate

Not applicable

## Consent for publication

Not applicable

